# A general framework to support cost-efficient survey design choices for the control of soil-transmitted helminths when deploying Kato-Katz thick smear

**DOI:** 10.1101/2023.02.13.23285837

**Authors:** Adama Kazienga, Bruno Levecke, Gemechu Tadesse Leta, Sake J. de Vlas, Luc E. Coffeng

## Abstract

**Background:** To monitor and evaluate soil-transmitted helminth (STH) control programs, the World Health Organization (WHO) recommends screening stools from 250 children across 5 schools, deploying Kato-Katz thick smear (KK). However, it remains unclear whether these recommendations are sufficient to make adequate decisions about stopping preventive chemotherapy (PC) (prevalence of infection <2%) or declaring elimination of STH as a public health problem (prevalence of moderate-to-heavy intensity (MHI) infections <2%).

**Methodology:** We developed a simulation framework to determine the effectiveness and cost of survey designs for decision-making in STH control programs, capturing the operational resources to perform surveys, the variation in egg counts across STH species, across schools, between and within individuals, and between repeated smears. Using this framework and a lot quality assurance sampling approach, we determined the most cost-efficient survey designs (number of schools, subjects, stool samples per subject, and smears per stool sample) for decision-making.

**Principal findings:** For all species, employing duplicate KK (sampling 4 to 6 schools and 64 to 70 subjects per school) was the most cost-efficient survey design to assess whether prevalence of any infection intensity was above or under 2%. For prevalence of MHI infections, single KK was the most cost-efficient (sampling 11 to 25 schools and 52 to 84 children per school).

**Conclusions/Significance:** KK is valuable for monitoring and evaluation of STH control programs, though we recommend to deploy a duplicate KK on a single stool sample to stop PC, and a single KK to declare the elimination of STH as a public health problem.

**Author summary:** Worldwide large-scale deworming programs are implemented to reduce the morbidity attributable to intestinal worms in school children. To monitor and evaluate the progress towards the programs goals, Word Health Organization (WHO) has developed both a survey design and a corresponding decision tree based on the prevalence and intensity of infections. However, these programs operate in resource-constrained countries, and hence it is crucial to minimize the operational costs to survey worm infections while ensuring the correctness of the program decision. To further support WHO in more evidence-based recommendations for cost-efficient decision-making, we developed a general framework that captures both the operational resources to perform surveys and the variation in test results when deploying the current diagnostic standard. Subsequently, we determined the most cost-efficient survey design to decide to stop the deworming programs and to verify whether the morbidity attributable to intestinal worms has been eliminated as a public health problem. Generally, we found that the current WHO-recommended survey design may not allow for optimal decision making. Based on our results, we proposed alternative survey designs for each of the worm species and program targets.

## Introduction

Recently, the World Health Organization (WHO) released its 2030 roadmap for the control of soil-transmitted helminths (STHs) [1,2]. In this new roadmap, the two crucial targets are (i) to eliminate STHs as a public health problem (EPHP), defined as prevalence of moderate-to-heavy intensity (MHI) infections less than 2%, and (ii) to reduce the number of tablets needed in preventive chemotherapy (PC). Regarding this second target, WHO recommends to stop PC if the prevalence of infection (any intensity) is less than 2%.

To assess whether prevalence of any intensity or MHI infection is under 2%, the WHO recommends screening stool samples from 250 children across 5 schools (50 children per school), using single Kato-Katz thick smears (KK) [2]. However, little attention has been paid to whether this survey design is sufficient for reliable decision-making. There will always be a risk of making a wrong decision: stopping PC or declaring EPHP too early (referred to as “undertreatment” from here on) or too late (“overtreatment”). Amongst other things, these risks are driven by survey sample size and diagnostic accuracy, which means that a trade-off has to be made between operational costs and feasibility of particular survey designs, and the maximum acceptable risks of under- and overtreatment.

Here, we assess the trade-off of survey design and associated operational costs versus the risk of under- and overtreatment for STHs when using KK as a diagnostic method because it was found more cost-effective than alternative diagnostic techniques based on egg counting [3]. We do this using a previously developed simulation framework for lot quality assurance sampling (LQAS) that only captured the prevalence of infection and not the variation in intensity of infection [4]. For the purpose of this study, we expanded this framework to also capture variation in intensity of infection across STH species (*Ascaris lumbricoides, Trichuris trichiura*, hookworms), across schools, between and within individuals, and between repeated thick smears. These sources of variation are important as the sensitivity of KK for detecting infection in an individual depends on the intensity of infection [5–9]. Furthermore, using this expanded framework, we assess to what extent the accuracy of survey results can be improved (and at what cost) by increasing the number of thick smears per stool sample or the number of stool samples per person [5,10]. Finally, for different levels of acceptable risk of under- and overtreatment, we determine the most cost-efficient survey design and the associated decision cut-off (maximum number of egg-positive individuals) for decision-making.

## Methods

### Overview

Based on a previously developed LQAS framework for monitoring and evaluation (M&E) of neglected tropical diseases (NTDs) [4], we developed a generic simulation framework for STH surveys and the associated operational costs at the level of a PC implementation unit. Using this framework, we assessed the cost-efficiency of different survey designs (number of schools, number of children per school, number of stool samples per child, and number of KK thick smears per stool sample) for each STH species and decision type (stop PC or declare EPHP) via the following steps: (1) simulate egg counts in thick smears of sampled children and determine the number of children that test positive; (2) for all possible decision cut-offs in terms of maximum number of positive children, calculate the probability of overtreatment and undertreatment across repeated Monte Carlo simulations and determine the decision cut-off for adequate decision-making (acceptable risk of under- and overtreatment); (3) estimate the total survey cost. To determine the most cost-efficient survey design and associated cut-off, we selected all designs that resulted in adequate decision-making and compared them in terms of cost and feasibility to implement.

### Simulation of egg counts for different survey designs

We adopted the simulation framework for egg counts developed by Coffeng et al. [11] and expanded it with geographical variation in infection levels. The expanded framework simulates egg count data from a compound lognormal-gamma-gamma-gamma-Poisson distribution, capturing the following sources of variability in egg counts:

1. Variability in mean egg per gram stool (EPG) between schools (assumed to be lognormal distributed (see **Supplement S1**);
2. Inter-individual variability in EPG due to variation in infection levels between individuals (considered to be gamma-distributed), where the level of aggregation (shape of the gamma distribution) is a linear function of the school-level mean EPG (see **Supplement S1**);
3. Day-to-day variability in mean EPG within an individual due to heterogeneous egg shedding over time (assumed to be gamma-distributed);
4. Variability in mean EPG between repeated slides based on the same sample due to the aggregated distribution of eggs in feces (assumed to follow a gamma distribution);
5. Variability in count observation (expressed in raw egg count) due to random diagnostic variation (assumed to be Poisson distributed).

We presented the mathematical backbone of the simulation framework in **Supplement S2** and quantified these aforementioned sources of variation based on published datasets [5,12]. **Table 1** provides the full overview of the estimated parameter values for each STH species.

**Table 1.** Parametrization of the simulation framework for various sources of variability in Kato-Katz thick smear egg counts. We parametrized the lognormal distribution using the mean and standard deviation on the logarithmic scale, and for the gamma distribution, the shape parameter *k* and rate 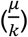, where *μ* is the distribution’s mean. To quantify *k*, the coefficient of variation (*cv*) was used as standardized measure of variability [13].

| Parameters | STH species |  |  | Data source |
| --- | --- | --- | --- | --- |
|  | *Ascaris* | Hookworm | *Trichuris* |  |
| Variability in mean EPG across schools within the same district ($\sigma_i$) | 0.69 | 0.80 | 0.52 | [12] |
| Intercept for school-level aggregation parameter ($\beta_0$) as a linear function of the school-level mean EPG | 0.0158 | 0.0162 | 0.0098 | [12] |
| Slope for school-level aggregation parameter ($\beta_1$) as a linear function of the school-level mean EPG | 0.0019 | 0.0222 | 0.0444 | [12] |
| Day-to-day variation in EPG within an individual (shape $k_d$, expressed as $\frac{1}{cv^2}$) | 0.5102 | 0.8734 | 1.4172 | [3] |

Using this simulation framework, we considered 4 survey designs, denoted as *KK*_*a* × *b*_, *a* is the number of stool samples per person, and *b* is the number of repeated smears per stool sample. These survey designs were *KK*_1 ×1_ (considered as the reference [1,2]), *KK*_1 × 2_, *KK*_2 × 1_, and *KK*_2 × 2_. We further set the number of schools (*n*_*schools*_) ranging from 3 to 10 for any intensity of infection and from 10 to 25 for MHI infections, while the number of children per school (*n*_*children*_) ranged from 10 to 200 for both targets.

### Calculating the probability of under- and overtreatment

To determine the probability of making right and wrong policy decisions for different survey designs, we used a 2-stage LQAS approach, a framework for NTD control program decision-making which we have described in detail previously [4]. The 2-stage aspect refers to the fact that we explicitly consider that survey results will be sampled from multiple clusters. In 2-stage LQAS, decision-making is based on the total number of positive test results (*X*_+_; here, egg-positive individuals) in a sample of pre-defined size from a pre-defined number of clusters in an implementation unit and a decision cut-off *c*. The frequency of PC will be reduced (or PC is stopped altogether) in case *X*_+_ is less than the decision cut-off *c*. Otherwise, the frequency of PC remains unchanged or may even be increased. This framework was used to assess two features of the decision: (1) the operational program decision threshold (e.g., 2% prevalence of infection); (2) the maximum acceptable risk of making a wrong decision when the true prevalence based on a single KK is at a pre-defined lower or higher value than the operational decision threshold (i.e., the “grey zone”, e.g., 1% and 3%). Using LQAS, we assessed two operational decision thresholds for each STH species: one for a 2% prevalence of any intensity of infection (i.e., related to stopping PC) and another for a 2% prevalence of MHI infections (i.e., related to declare EPHP). Then, we considered adequate decision-making by setting the maximum allowed probability of unnecessarily continuing with the program (*E*_*overtreat*_) when true prevalence is 3% (the upper limit of the grey zone, or *UL*) at 0.25. We further set the highest allowed probability of prematurely stopping interventions (*E*_*undertreat*_) when true prevalence is 1% (the lower limit of the grey zone, or *LL*) at 0.05. For this, we determined the mean EPG at the implementation unit level that corresponds to these limits (*LL* and *UL*) as measured by single KK (**Table 2**), conditional on the estimated parameter values for sources of variability in egg counts (**Table 1**), assuming 100% specificity of KK [14,15].

**Table 2.** Parametrization of simulations for 2-stage lot quality assurance sampling in terms of the natural logarithm of the geometric mean infection intensity (eggs per gram feces or EPG) in the population as measured by single Kato-Katz.

| Parameters | STH species |  |  |
| --- | --- | --- | --- |
|  | *Ascaris* | Hookworm | *Trichuris* |
| ***Any intensity infections*** |  |  |  |
| Lower limit mean EPG | log(0.449) | log(0.280) | log(0.298) |
| Upper limit mean EPG | log(2.691) | log(1.013) | log(0.980) |
| ***Moderate-to-heavy intensity infections*** |  |  |  |
| Lower limit mean EPG | log(314) | log(164) | log(145) |
| Upper limit mean EPG | log(620) | log(287) | log(217) |

### Calculating the operational costs and determining the most cost-efficient survey design

We estimated the total survey costs for each simulated survey design and STH species. This cost was composed of (*i*) the cost of consumables to collect and process samples, (*ii*) the personnel costs, and (*iii*) the travel costs [4].Technical details on how the total survey costs were estimated can be found in **Supplement S3**.

To determine the most cost-efficient survey design and associated decision cut-off, we selected all designs that allowed for adequate decision-making. Then, we chose the survey design that resulted in the lowest total survey cost (*C*_*tot*_) and required a maximum of 100 children per school (a reasonable maximum size of individuals in a school).

### Sensitivity analysis

We assessed the impact of several critical parameters on the estimated cost-efficiency of different survey designs. First, we explored the impact of assuming that variation between individuals within schools is governed by a fixed aggregation parameter *k*_*k*_ (in contrast to letting it vary as a linear function of the mean EPG in a school). Second, we assessed the impact of considering higher inter-individual variability in egg counts (*k*_*k*_) in (post-) control scenarios due to incomplete deworming coverage compared to this variability in the main analysis (assumed to be as observed in pre-control settings). Therefore, we set *k*_*k*_ to 2/3 of the value in the main analysis. Third, because diagnostic performance impacts survey design [4,16], we evaluated the impact of KK specificity, assuming 99.5% specificity instead of 100% in the main analysis. Finally, to assess the impact of stricter choices about the maximum allowed probability of making incorrect decisions, we set the risk of undertreating (*E*_*undertreat*_) to 0.025 instead of 0.05 in the main analysis. As a second step, we also decreased the risk of overtreating (*E*_*overtreat*_) from 0.25 to 0.1.

## Results

### Cost-efficient survey design for stopping PC

**Fig 1** illustrates the required number of children per school (*n*_*children*_) and the total survey cost (*C*_*tot*_) as a function of the number of sampled schools (*n*_*schools*_) to reliably stop PC for the different survey designs for each of the three STH species separately. The panels on *n*_*children*_ (**Fig 1A, 1C** and **1E**) highlight three important aspects. First, fewer children per school are required when more schools are included in the survey. Second, preparing more smears per sample and collecting more stools per child further reduces the number of children per school. For example, when making program decisions for *Ascaris* infections and sampling 5 schools (**Fig 1A**), 88 children per school are required for a *KK*_1 × 1_ survey design, while this number reduces to 64, 56, and 52 when deploying *KK*_1 × 2_, *KK*_2 × 1_ and *KK*_2 × 2_ survey designs, respectively. Third, the minimum required size of the surveys varies across the three STH species. For instance, when sampling 5 schools and deploying a *KK*_1 × 1_ survey design, 88 children per school are required for *Ascaris* infections (**Fig 1A**), while this is 126 for hookworm infections (**Fig 1C**) and 80 for *Trichuris* (**Fig 1C**).

**Fig 1.**
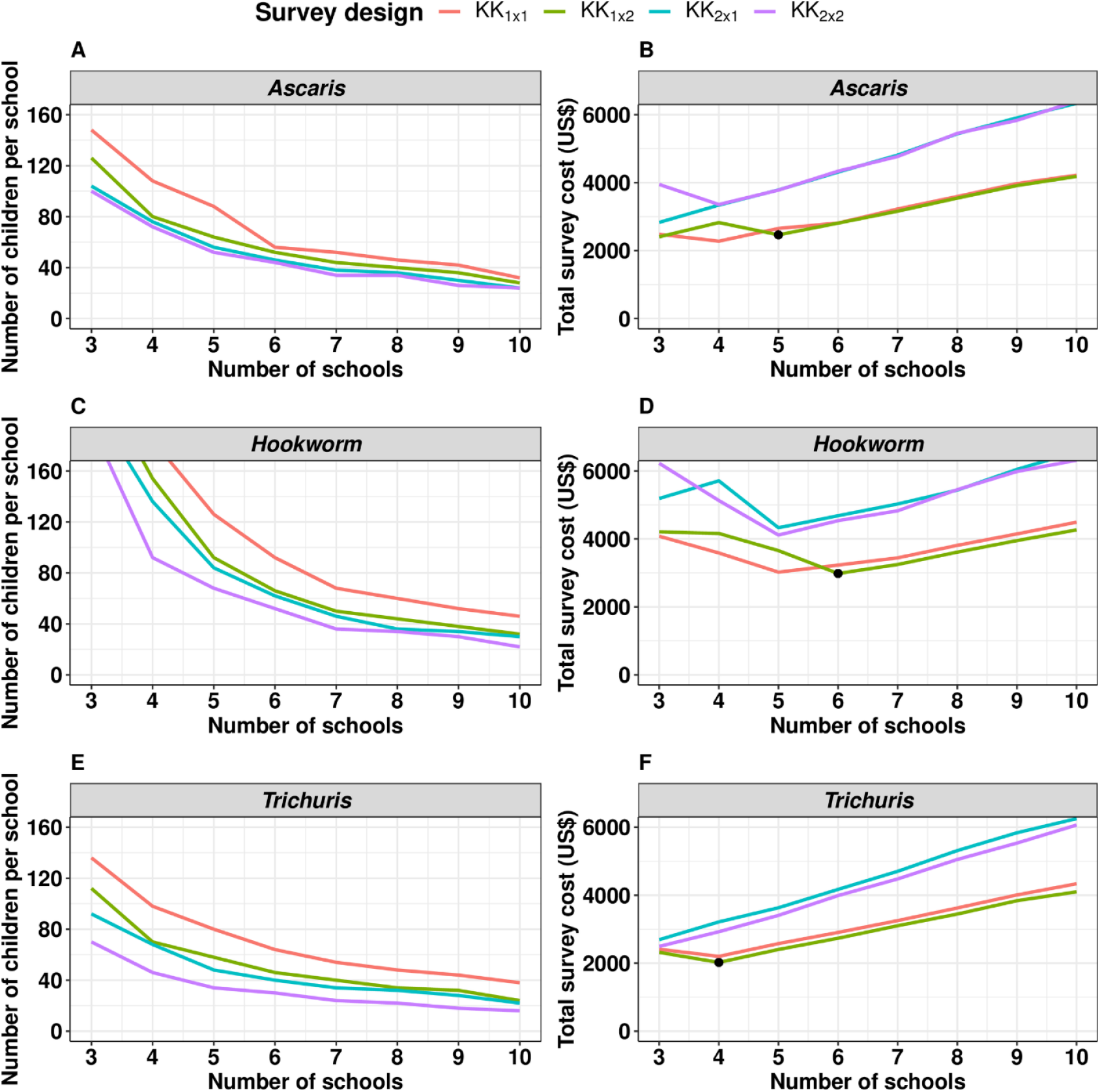
The identification of the most cost-efficient survey design to reliably stop preventive chemotherapy. This figure illustrates the required number of children per school (*n*_*children*_; **Panels A, C** and **E**) and the corresponding total survey cost (*C*_*tot*_; **Panels B, D** and **F**) as a function of the number of sampled schools (*n*_*schools*_) for different survey designs and soil-transmitted helminths (*Ascaris*: **Panels A** and **B**; hookworm: **Panels C** and **D**; *Trichuris*: **Panels E** and **F**). The survey designs (*KK*_*a* × *b*_) varied in the number of stool samples per child (*= a*) and the number of Kato-Katz thick smears per sample (*= b*). The black bullet point in Panels B, D and F indicates the most cost-efficient survey design. In other words, the number of schools that minimizes the costs while ensuring reliable decision-making. Note that we assumed that the maximum number of children per school could not exceed 100.

When focusing on the panels representing the total cost *C*_*tot*_ (**Fig 1B, 1D** and **1F**), two additional aspects become apparent. Survey designs involving a second stool (*KK*_2 × 1_ and *KK*_2 × 2_) are more financially demanding, even though fewer children are required to be sampled. Re-taking the aforementioned example (making program decisions for *Ascaris* infections and sampling 5 schools), the total survey costs for *KK*_1 × 1_ survey design equals 2,653 US$ (*n*_*children*_ = 88), while survey costs are up to 3,781 US$ when deploying a *KK*_2 × 2_ survey design (*n*_*children*_ = 52). Second, after an initial drop, the total survey costs further increase as a function of number of schools (for *KK*_1 × 1_ and *KK*_1 × 2_ survey designs across all STH species). In other words, there is an optimal number of schools that minimizes the costs while ensuring reliable decision-making.

Combining all these aspects and considering that some schools do not have more than 100 children, *KK*_1 × 2_ is the most cost-efficient of all feasible survey designs for all three STH species, but the difference with *KK*_1 × 1_ is modest. However, the required sample size (*n*_*schools*_ × *n*_*children*_) and the total survey costs varies across the 3 STH species (*Ascaris*: 5 × 64, 2,465 US$ (**Fig 1B**); hookworm: 6 × 66, 2,983 US$ (**Fig 1D**); *Trichuris*: 4 × 70, 2,022 US$ (**Fig 1F**)).

In **Table 3**, we explored the impact of assumptions about inter-individual variation in EPG and the specificity of KK on what is the optimal survey design (*n*_*schools*_ *x n*_*children*_) for the different STH species separately. This table indicates three important patterns when considering *Ascaris*. First, compared to an aggregation of infections within children (*k*_*k*_), varying as a function of school mean EPG (5 × 64), fixing *k*_*k*_ (= 1.75) resulted in a smaller sample size (3 × 90) and a reduction of the total survey costs of 11.5% (= 2,181 US$/ 2,465 US$). Second, when assuming a post-control *k*_*k*_ (= 2/3 x varying *k*_*k*_) resulted in slight increase in both sample size (5 × 68) and total survey costs (1.7% = 2,507 US$/ 2,465 US$). Finally, a reduced clinical specificity (= 99.5%) increased the sample size from 5 × 64 to 7 × 62, requiring 38.8% more funds. These patterns were very similar for the other species (**Table 3**).

**Table 3.** The impact of alternative model assumptions on both the required sample size and the total survey costs to reliably stop preventive chemotherapy. This table presents the sample size (*n*_*schools*_ × *n*_*children*_) the decision cut-off *c* and the associated total survey costs (*C*_*tot*_) for different scenarios of parameterizing the aggregation of infections within children (*k*_*k*_), the clinical specificity of Kato-Katz thick smear and the maximum allowed risk of incorrect decision-making for the different soil-transmitted helminths species separately. As a reference we assumed that *k*_*k*_ varies as a function of school mean eggs per gram of stool (EPG) and that Kato-Katz thick smears has a perfect specificity. We allowed for a risk of undertreatment (*E*_*undertreat*_) equal to 0.05 and risk of overtreatment (*E*_*overtreat*_) equal to 0.25. Also, the fixed aggregation parameter (Fixed *k*_*k*_), which captures the variation between individuals within schools, was 0.326 for *Ascaris*, 0.257 for hookworm, and 0.532 for *Trichuris*. A lower value of *k*_*k*_ (Lower *k*_*k*_) means higher inter-individual variation in EPG values, assumed to be 2/3 of *k*_*k*_ in the main analysis.

| Parameters | *Ascaris* |  |  | Hookworm |  |  | *Trichuris* |  |  |
| --- | --- | --- | --- | --- | --- | --- | --- | --- | --- |
| | Sample size | $c$ | $C_{tot}$ (US\$) | Sample size | $c$ | $C_{tot}$ (US\$) | Sample size | $c$ | $C_{tot}$ (US\$) |
| **Aggregation of infections within children ($k_k$)** |  |  |  |  |  |  |  |  |  |
| Varying $k_k$ as a linear function of school mean EPG | 5 x 64 | 7 | 2,465 | 6 x 66 | 9 | 2,983 | 4 x 70 | 7 | 2,022 |
| Fixed $k_k$ | 3 x 90 | 7 | 2,181 | 6 x 58 | 9 | 2,883 | 3 x 72 | 6 | 1,529 |
| Lower $k_k$ | 5 x 68 | 7 | 2,507 | 6 x 70 | 9 | 3,033 | 5 x 58 | 7 | 2,403 |
| | Sample size | $c$ | $C_{tot}$ (US\$) | Sample size | $c$ | $C_{tot}$ (US\$) | Sample size | $c$ | $C_{tot}$ (US\$) |
| **Specificity of Kato-Katz thick smear** |  |  |  |  |  |  |  |  |  |
| 100% | 5 x 64 | 7 | 2,465 | 6 x 66 | 9 | 2,983 | 4 x 70 | 7 | 2,022 |
| 99.5% | 7 x 62 | 12 | 3,422 | 8 x 70 | 17 | 4,044 | 6 x 58 | 11 | 2,883 |
| **Risks of incorrect decision-making** |  |  |  |  |  |  |  |  |  |
| $E_{undertreat} = 0.05; E_{overtreat} = 0.25$ | 5 x 64 | 7 | 2,465 | 6 x 66 | 9 | 2,983 | 4 x 70 | 7 | 2,022 |
| $E_{undertreat} = 0.025; E_{overtreat} = 0.25$ | 6 x 72 | 9 | 3,058 | 8 x 56 | 10 | 3,811 | 5 x 68 | 8 | 2,507 |
| $E_{undertreat} = 0.025; E_{overtreat} = 0.10$ | 9 x 62 | 13 | 4,400 | 10 x 72 | 18 | 5,097 | 8 x 62 | 13 | 3,911 |

We also verified the additional survey costs to further minimize the probability of over- and undertreatment (**Table 3**). Obviously, further minimizing the risk of undertreatment (*E*_*undertreat*_) and overtreatment (*E*_*overtreat*_) increases the sample size and hence the total survey cost, but not extremely. When we reduce the maximum allowed risk of undertreatment from 0.05 to 0.025, the total survey costs increase by 24% (3,058 US$ *vs*. 2,465 US$). When we also reduce the risk of undertreatment from 0.25 to 0.10, the sample size further increases to 78.5% (4,400 US$ *vs*. 2,465 US$). Again, we observed similar patterns for the other two species (**Table 3**).

### Cost-efficient survey design to declare elimination of STH as a public health problem

After having assessed study designs and decision cut-offs for stopping PC, we now turn to study designs for declaring elimination of STH as a public health problem (**Table 4; Supplement S4, Figure S1**). Compared to the most cost-effective survey designs for stopping PC, two important differences can be noted. First, the *KK*_1 × 1_ rather than *KK*_1 × 2_ survey design is the most cost-efficient choice. Second, the required sample size (*n*_*schools*_ × *n*_*children*_) is substantially larger (*Ascaris*: 11 × 84 *vs*. 5 × 64; hookworm: 25 × 52 *vs*. 6 × 66; *Trichuris*: 19 × 68 *vs*. 4 × 70). Compared to the total survey costs to reliably stop PC, the cost of surveys to reliably declare EPHP are 2.3 (*Ascaris*) to 4.6 (*Trichuris*) times higher.

**Table 4.** The cost-efficient survey design to declare elimination of soil-transmitted helminths as a public health problem. This table represents the required sample size (*n*_*schools*_ × *n*_*children*_), the decision cut-off *c* and the total survey costs (*C*_*tot*_) for four different survey designs (*KK*_*a* × *b*_) that vary in the number of stool samples per child (*= a*) and the number of Kato-Katz thick smears per sample (*= b*)

| STH | Survey design | $n_{schools} \times n_{children}$ | $c$ | $C_{tot}$ (US\$) |
| --- | --- | --- | --- | --- |
| *Ascaris* | $KK_{1 \times 1}$ | 11 x 84 | 13 | 5,775 |
| Hookworm | $KK_{1 \times 1}$ | 25 x 52 | 17 | 11,522 |
| *Trichuris* | $KK_{1 \times 1}$ | 19 x 68 | 16 | 9,346 |

We also explored the impact of the assumptions made on the parameterization of the aggregation in infections between children (*k*_*k*_), the clinical specificity and the allowed risk for both over- and undertreatment on the sample size (*n*_*schools*_ × *n*_*children*_) and the total survey cost. Not unexpectedly, we observed similar trends as for reliably stopping PC, and hence we refer to **Supplement S4, Table S1** for more details.

## Discussion

We have demonstrated that the current WHO guidelines to M&E STH control programs may not guarantee reliable decision-making for all program targets and STH species. Based on our results, we rather recommend deploying a duplicate KK on one stool when making decisions to stop PC, deploying a single KK on one stool sample when the aim is to verify whether STH has been eliminated as a public health problem. This difference in survey design is in line with other researchers’ findings [10] and can be explained by the fact that examining multiple KK smears increases the sensitivity [5,10,17,18] but generally does not affect estimates of the mean intensity of infection [5,10,17,19]. Also, collecting samples on two consecutive days (*KK*_2 × 1_ and *KK*_2 × 2_) was not cost-efficient, because it requires more funds. In addition, as the required sample size varies across STH species (due to differences in fecundity and aggregation of infections; see **Table 1**), it will be important to ensure sufficient number of schools and children per school to arrive at reliable decision-making for all STH present in the surveyed implementation unit. In other words, the sample size is dictated by the STH species that requires the largest number of schools and children. This means that in an area where all three STH are present, the sample size to reliably stop PC is dictated by hookworms (6 schools and 66 children per school; see **Table 3**). Note that STH-specific decision cut-offs vary across sample size, and hence they will need to be adapted accordingly. In **Supplement S4, Tables S2** and **S3**, we therefore provide the required sample size and the corresponding STH-specific decision cut-offs for areas where more than STH species are considered for decision-making.

We showed that the aggregation of infections within children (*k*_*k*_) varies as a function of school mean EPG (see **Supplement S1**), which is in agreement with a recent analysis of the TUMIKIA data. Indeed, this analysis showed that if not considered properly, it leads to poor decision-making due to smaller-than-required sample sizes [20]. Furthermore, the impact of reduced clinical specificity (resulting in increased sample size and total survey cost) was in line with previous work [4], highlighting the need for highly specific diagnostic methods when targeting low-intensity infections settings [21–23].

Minimizing the risk of making incorrect program decisions (risk of overtreatment and undertreatment) is more financially demanding. However, by improving decision-making, one will also save substantial costs due to unnecessarily distributing tablets or re-establishing the PC program. Furthermore, future research using transmission models is needed to assess the appropriateness of the WHO-recommended thresholds for STH policy.

The LQAS approach in this simulation study has also been used for population-based decision-making in schistosomiasis [24] and trachoma [25] control programs. Conversely, another approach is the model-based geostatistical methods for M&E of STH, and other NTDs [26,27]. Yet, compared to the LQAS method, this alternative approach adoption required additional expertise in geostatistical analysis [4,27].

## Conclusion

We confirm that KK is valuable for M&E of STH in low-endemicity infections settings, but the current WHO-recommended survey design may not guarantee reliable decision-making to stop PC, and especially not for declaring EPHP. We, therefore, provide cost-efficient alternative survey designs.

## Data Availability

Yes - all data are fully available without restriction

## Supporting Information Legends

**Supplementary information S1**. Quantification of between and within -school variation in faecal egg counts

**Supplementary information S2**. The mathematical backbone of the simulation framework

**Supplementary information S3**. Estimation of total survey cost to monitor and evaluate STH when using Kato-Katz thick smear

**Supplementary information S4**. Additional tables and figures

## References

1. World Health Organization. Ending the neglect to attain the Sustainable Development Goals: A road map for neglected tropical diseases 2021–2030. In: World Health Organization [Internet]. 2021 [cited 10 Feb 2022]. Available: https://www.who.int/publications/i/item/9789240010352

2. World Health Organization. 2030 targets for soil-transmitted helminthiases control programmes. World Health Organization; 2020 [cited 15 Mar 2022]. Available: https://apps.who.int/iris/handle/10665/330611

3. Coffeng LE, Vlaminck J, Cools P, Denwood M, Albonico M, Ame SM, et al. A general framework to support cost-efficient fecal egg count method and study design choices for large-scale STH deworming programs – monitoring of therapeutic drug efficacy as a case study. medRxiv. 2023; 2023.01.06.23284253. doi:10.1101/2023.01.06.23284253

4. Kazienga A, Coffeng LE, de Vlas SJ, Levecke B. Two-stage lot quality assurance sampling framework for monitoring and evaluation of neglected tropical diseases, allowing for imperfect diagnostics and spatial heterogeneity. PLoS Negl Trop Dis. 2022;16: e0010353.

5. Cools P, Vlaminck J, Albonico M, Ame S, Ayana M, José Antonio BP, et al. Diagnostic performance of a single and duplicate Kato-Katz, Mini-FLOTAC, FECPAKG2 and qPCR for the detection and quantification of soil-transmitted helminths in three endemic countries. PLoS Negl Trop Dis. 2019;13: e0007446.

6. Khurana S, Sethi S. Laboratory diagnosis of soil transmitted helminthiasis. Trop Parasitol. 2017;7: 86.

7. Niguse AF, Hailu T, Alemu M, Nibret E, Amor A, Munshea A. Evaluating the Performance of Diagnostic methods for soil transmitted helminths in the Amhara National Regional State, Northwest Ethiopia. BMC Infect Dis. 2020.

8. Dunn JC, Papaiakovou M, Han KT, Chooneea D, Bettis AA, Wyine NY, et al. The increased sensitivity of qPCR in comparison to Kato-Katz is required for the accurate assessment of the prevalence of soil-transmitted helminth infection in settings that have received multiple rounds of mass drug administration. Parasit Vectors. 2020;13: 1–11.

9. De Vlas SJ, Gryseels B. Underestimation of Schistosoma mansoni prevalences. Parasitol Today. 1992;8: 274–277.

10. Coffeng LE, Malizia V, Vegvari C, Cools P, Halliday KE, Levecke B, et al. Impact of different sampling schemes for decision making in soil-transmitted helminthiasis control programs. J Infect Dis. 2020;221: S531–S538.

11. Coffeng LE, Levecke B, Hattendorf J, Walker M, Denwood MJ. Survey design to monitor drug efficacy for the control of soil-transmitted helminthiasis and schistosomiasis. Clin Infect Dis. 2021;72: S195–S202.

12. Leta GT, Mekete K, Wuletaw Y, Gebretsadik A, Sime H, Mekasha S, et al. National mapping of soil-transmitted helminth and schistosome infections in Ethiopia. Parasit Vectors. 2020;13: 1–13.

13. Denwood MJ, Love S, Innocent GT, Matthews L, McKendrick IJ, Hillary N, et al. Quantifying the sources of variability in equine faecal egg counts: implications for improving the utility of the method. Vet Parasitol. 2012;188: 120–126.

14. Speich B, Ali SM, Ame SM, Albonico M, Utzinger J, Keiser J. Quality control in the diagnosis of Trichuris trichiura and Ascaris lumbricoides using the Kato-Katz technique: experience from three randomised controlled trials. Parasit Vectors. 2015;8: 1–8.

15. Knopp S, Salim N, Schindler T, Voules DAK, Rothen J, Lweno O, et al. Diagnostic accuracy of Kato–Katz, FLOTAC, Baermann, and PCR methods for the detection of light-intensity hookworm and Strongyloides stercoralis infections in Tanzania. Am J Trop Med Hyg. 2014;90: 535.

16. Levecke B, Coffeng LE, Hanna C, Pullan RL, Gass KM. Assessment of the required performance and the development of corresponding program decision rules for neglected tropical diseases diagnostic tests: Monitoring and evaluation of soil-transmitted helminthiasis control programs as a case study. PLoS Negl Trop Dis. 2021;15: e0009740.

17. Moser W, Bärenbold O, Mirams GJ, Cools P, Vlaminck J, Ali SM, et al. Diagnostic comparison between FECPAKG2 and the Kato-Katz method for analyzing soil-transmitted helminth eggs in stool. PLoS Negl Trop Dis. 2018;12: e0006562.

18. Nikolay B, Brooker SJ, Pullan RL. Sensitivity of diagnostic tests for human soil-transmitted helminth infections: a meta-analysis in the absence of a true gold standard. Int J Parasitol. 2014;44: 765–774.

19. Levecke B, Brooker SJ, Knopp S, Steinmann P, Sousa-Figueiredo JC, Stothard JR, et al. Effect of sampling and diagnostic effort on the assessment of schistosomiasis and soil-transmitted helminthiasis and drug efficacy: a meta-analysis of six drug efficacy trials and one epidemiological survey. Parasitology. 2014;141: 1826–1840.

20. Truscott JE, Ower AK, Werkman M, Halliday K, Oswald WE, Gichuki PM, et al. Heterogeneity in transmission parameters of hookworm infection within the baseline data from the TUMIKIA study in Kenya. Parasit Vectors. 2019;12: 1– 13.

21. Diagnostic target product profiles for monitoring, evaluation and surveillance of schistosomiasis control programmes. [cited 11 Oct 2021]. Available: https://www.who.int/publications/i/item/9789240031104

22. Stuyver LJ, Levecke B. The role of diagnostic technologies to measure progress toward WHO 2030 targets for soil-transmitted helminth control programs. PLoS Negl Trop Dis. 2021;15: e0009422.

23. Lim MD, Brooker SJ, Belizario Jr VY, Gay-Andrieu F, Gilleard J, Levecke B, et al. Diagnostic tools for soil-transmitted helminths control and elimination programs: A pathway for diagnostic product development. Public Library of Science San Francisco, CA USA; 2018.

24. Olives C, Valadez JJ, Brooker SJ, Pagano M. Multiple category-lot quality assurance sampling: a new classification system with application to schistosomiasis control. PLoS Negl Trop Dis. 2012;6: e1806.

25. Myatt M, Mai NP, Quynh NQ, Nga NH, Tai HH, Long NH, et al. Using lot quality-assurance sampling and area sampling to identify priority areas for trachoma control: Viet Nam. Bull World Health Organ. 2005;83: 756–763.

26. Diggle PJ, Amoah B, Fronterre C, Giorgi E, Johnson O. Rethinking neglected tropical disease prevalence survey design and analysis: a geospatial paradigm. Trans R Soc Trop Med Hyg. 2021;115: 208–210.

27. Johnson O, Fronterre C, Amoah B, Montresor A, Giorgi E, Midzi N, et al. Model-based geostatistical methods enable efficient design and analysis of prevalence surveys for soil-transmitted helminth infection and other neglected tropical diseases. Clin Infect Dis. 2021;72: S172–S179.

